# Within-visit Blood Pressure Variability in Children and Adolescents in the National Health and Nutrition Examination Survey (2013-2020)

**DOI:** 10.1101/2024.08.14.24312023

**Authors:** Sandeep K Riar, Scott Gillespie, Andrew M South

## Abstract

**Background:** It is assumed that BP decreases with repeat measurements and multiple readings are recommended. There is limited information about within-visit BP variability (BPV) in healthy children.

**Methods:** We used NHANES data (2013-2020) to measure BPV in subjects 8-17 years old with three BP readings. During 2013 to 2016, auscultatory BP was obtained (manual protocol, MP). Subsequently, oscillometric BP was measured (automated protocol, AP). We excluded subjects with diastolic BP ‘zero’.

**Results:** We included a total 5656 subjects with 3365 (59.5%) in the MP and 2291 (40.5%) in the AP group. A ΔBP (individual-level difference between the highest and lowest of three BP readings) ≥5 mmHg was noted in 49.1% and 60.7% subjects for systolic BP (SBP) and diastolic BP (DBP) respectively. A ΔDBP ≥10 mmHg and DBP average real variability ≥10 mmHg was twice as common in the MP than AP group. A ΔBP ≥20 mmHg was noted in 1.3% and 4.4% subjects for SBP and DBP respectively. The first SBP and DBP reading was ≥5 mmHg higher or lower than the averaged second and third SBP and DBP readings in 24.9% and 34.5% of subjects respectively. The first SBP and DBP reading was the highest of three BP readings in 44.2% and 42.4% subjects respectively.

**Conclusions:** There is significant BPV in children and adolescents. Diastolic BPV is more common in MP than AP groups. Initial BP is not always the highest and inclusion of second and third BP reading may be more representative of patient’s actual BP.

## INTRODUCTION

Hypertension is a major cause of premature death worldwide, and children with high blood pressure (BP) have increased risk of elevated BP in adulthood.^1,2^ Accurate BP measurement is critical as a 5-mmHg difference in systolic BP (SBP) could lead to hypertension misdiagnosis in 84 million people worldwide.^3^ The deleterious impact of erroneous BP measurement is recognized, however even an accurate single BP reading is inadequate to diagnose hypertension.^4^ BP is a continuous variable that changes in response to endogenous factors and exogenous stimuli.^3^ Physiologic BP variability (BPV) in normotensive individuals allows for homeostasis.^5^ Multiple BP readings are recommended at each encounter as it is widely believed that the first BP reading will be higher than subsequent BP readings from ‘stress response/alerting reaction’, accommodation effect, regression to the mean, and higher inflation pressure with the first automated BP measurement.^6–10^

However, guidelines vary in the recommended number of BP readings and inclusion of the first reading. While adult guidelines recommend the average of 2–3 BP readings at each visit, pediatric hypertension diagnosis requires averaging the 2^nd^ and 3^rd^ BP readings and some use the lowest of three BP measurements.^11–14^ In real life, typically a single BP reading is obtained, likely from time and staff constraints and perhaps limited awareness of guidelines.^15–18^ The failure to obtain multiple BP readings may have significant implications. In adults, initial SBP reading was 12.7 mmHg higher compared to the average of three SBP readings.^15^ Notably, in adults and adolescents, the threshold BP for hypertension is 10 mmHg and 1 mmHg higher than normal SBP and diastolic BP (DBP) respectively (130 vs. 120 mmHg for SBP; ≥80 vs. <80 mmHg for DBP).^11,19^ However, in younger children, the difference is only about 4 mmHg and 3 mmHg respectively (95^th^ percentile vs. 90^th^ percentile).^11,20^ International BP device validation guidelines allow a maximum 5-mmHg difference between test and reference BP readings as one of the criteria.^10^ Office BP monitoring is indispensable to hypertension management and available studies of short-term BPV describe its impact on hypertension diagnosis but there is scant data on individual level within-visit BPV in healthy children. As compared to one BP reading, the use of two or three BP readings reduced BP classification in 6-9% of pediatric patients and 34% of adults.^14,21,22^ A study of within-visit BPV in Australian children noted mean SBP change of 6.7 mmHg between successive BP readings; however, the study included only two automated BP readings in the majority.^23^

The National Health and Nutrition Examination Survey (NHANES) examines a nationally representative sample of about 5,000 civilian, noninstitutionalized U.S. persons each year and is conducted by the National Center for Health Statistics. The BP protocol stipulates obtaining three BP readings within a single encounter for all participants 8 years and older.^24^ The objective of our study was to use pediatric data from NHANES as a convenience sample to: (1) describe variability of three BP readings; (2) compare the initial BP reading with the average of second and third BP readings and the average of all three BP readings; and (3) compare whether each of these measures vary by manual (auscultatory) vs. automated (oscillometric) BP readings.

## METHODS

The authors declare that all supporting data are available within the article and its online supplementary files. Additional data that support the findings of this study are available from the corresponding author upon reasonable request.

### Study design, BP measurement and terminology

NHANES is a cross-sectional survey that operates continuously, and data are released in 2-year cycles. We used data from three cycles (2013–2014, 2015–2016, and 2017–March 2020, the pre-pandemic cycle). Due to the COVID-19 pandemic, data for the NHANES 2019-2020 cycle was incomplete and not nationally representative and was combined with data from the NHANES 2017-2018 cycle to form a nationally representative sample.^25^ Study population included youth aged 8 to 17 years with three BP readings. NHANES was approved by the National Center for Health Statistics Research Ethics Review Board. Parental consent and participant assent were obtained.^26^

Demographic information (age, sex, race, and Hispanic origin) were collected during an interview.^27^ Standing height and weight were measured using standard methods and equipment. Body mass index (BMI) percentile was determined from the Centers for Disease Control and Prevention 2000 growth charts.^28^ Weight status was determined by BMI percentile and categorized dichotomously as underweight/healthy weight (BMI <85^th^ percentile) versus overweight/obesity (BMI ≥85^th^ percentile).

The BP methodology changed during the study period. During the 2013–2014 and 2015–2016 cycles, BP was measured using auscultation, referred to as the manual protocol (MP) here.^29,30^ In 2017 NHANES added oscillometric measurement of BP using an automated protocol (AP). In the 2017–2018 cycle, BP was measured using both MP and AP in each participant and since 2019 only AP is being used.^24,30,31^ To avoid cohort overlap, in this analysis we used only automated readings from the 2017–2018 cycle which are available in the 2017– March 2020 cycle. We used unweighted data as our goal was to use the NHANES database as a source of standardized BP readings in a large group of children and not to make estimates representative of the U.S. population.

NHANES used a standardized BP measurement protocol, with seated rest for five minutes prior to BP measurement.^24,29,30^ Right arm was preferred for BP measurement, but left arm could be used if needed. With the MP, a physician obtained three readings thirty seconds apart using a stethoscope and wall-mounted mercury sphygmomanometer.^29,30^ DBP was recorded as the pressure at which the last muffled sound was heard, also referred to as the fifth Korotkoff sound.^32^ The AP, performed by a technician, utilized the Omron HEM-907XL device that automatically obtained three BP measurements one minute apart.^24,30,31^ To minimize data entry error, NHANES has hard edits programmed into the system for each protocol. For MP: 1) SBP cannot be above 300 mmHg; 2) SBP must be above DBP; and 3) if there is no SBP then there cannot be a DBP although there can be an SBP without a DBP because SBP cannot be zero, but DBP can be zero.^33^ Similar hard edits are programmed into the system for AP, however neither SBP nor DBP can be zero.^24^

As studying BPV was an objective of our study, we did not perform data clean-up procedures for those with excessive differences among BP readings. We explored subjects with a DBP of zero but omitted them from data analysis for BPV and BP classification.

Hypertension-range BP readings were defined using the recommendations in the 2017 pediatric hypertension guidelines.^11^ Similar to other studies that used NHANES data to study pediatric hypertension, we used the average of all three BP readings for each subject to classify BP as hypertension range.^34,35^ Specifically, for children aged 8 to 12 years, we determined the BP percentile using age, sex, and height and defined hypertension as SBP and/or DBP ≥95^th^ percentile or SBP ≥130 and/or DBP ≥80 mmHg (whichever was lower). For age 13 to 17 years, hypertension was defined by BP ≥130/≥80 mmHg.

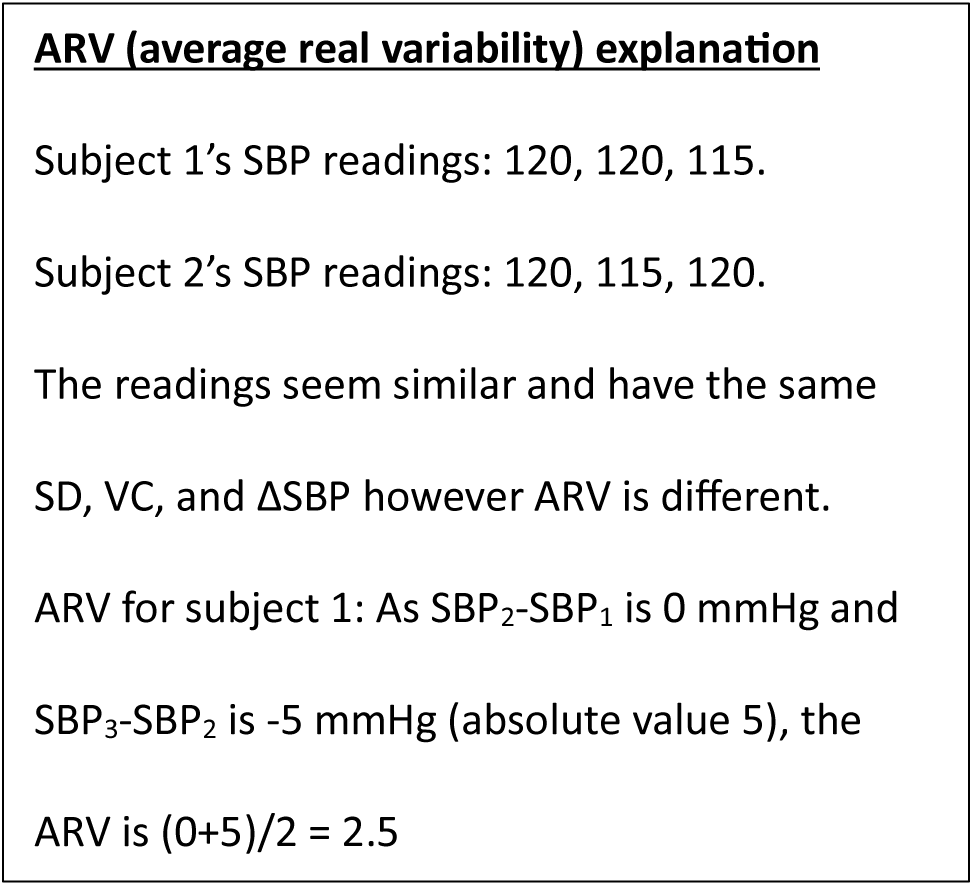

We assessed BPV among the three readings for each subject using measures that either ignored or considered the temporal order of BP readings, denoted as non-directional or directional respectively. Non-directional BPV measures included standard deviation (SD), variability coefficient (VC), and the difference between the highest and lowest BP reading for each subject (ΔBP). Directional BPV measures included average real variability (ARV), comparison, and trend among the three sequential BP readings. This also included comparison of the first BP reading (BP_1_) with the average of the next two BP readings (BP_2+3_, referred to as ’subsequent’ BP) as well as the average of all three BP readings (BP_1+2+3_, referred to as ‘total’ BP). The difference between the first BP reading and subsequent BP is labeled ΔBP_sub_, referred to as “white coat effect” (WCE) by some.^36^ The difference between the initial BP reading and total BP is referred to as ΔBP_total_. ARV is the average of the “absolute” differences between consecutive BP readings while accounting for the temporal order of BP measurements (see Box).^37^

### Statistical analysis

Statistical summaries for demographics, BP readings, and BPV were calculated using medians and interquartile ranges for continuous variables and frequencies and percentages for categorical data. Means and SD were additionally included in the display tables for some continuous variable analyses to allow for easy comparison with other studies. Comparisons between manual and automated BP readings were performed using Wilcoxon rank-sum tests, and comparisons between categorical data were performed using Chi-square test or Fisher exact test. All analyses were performed in SAS v.9.4 (Cary, NC), and statistical significance was assessed two-sided at the 0.05 threshold. No sample weights were incorporated in the analysis.

One of the authors (SR) had full access to all the data in the study and takes responsibility for its integrity and the data analysis.

## Results

Of the 6801 eligible subjects, 5874 (86.4%) had three BP readings (Figure 1). An additional 3.7% (218/5874) subjects were excluded due to one or more DBP readings of ‘zero’. We included a total of 5656 subjects in the primary analysis with 3365 (59.5%) in the MP and 2291 (40.5%) in the AP group.

**Figure 1:**
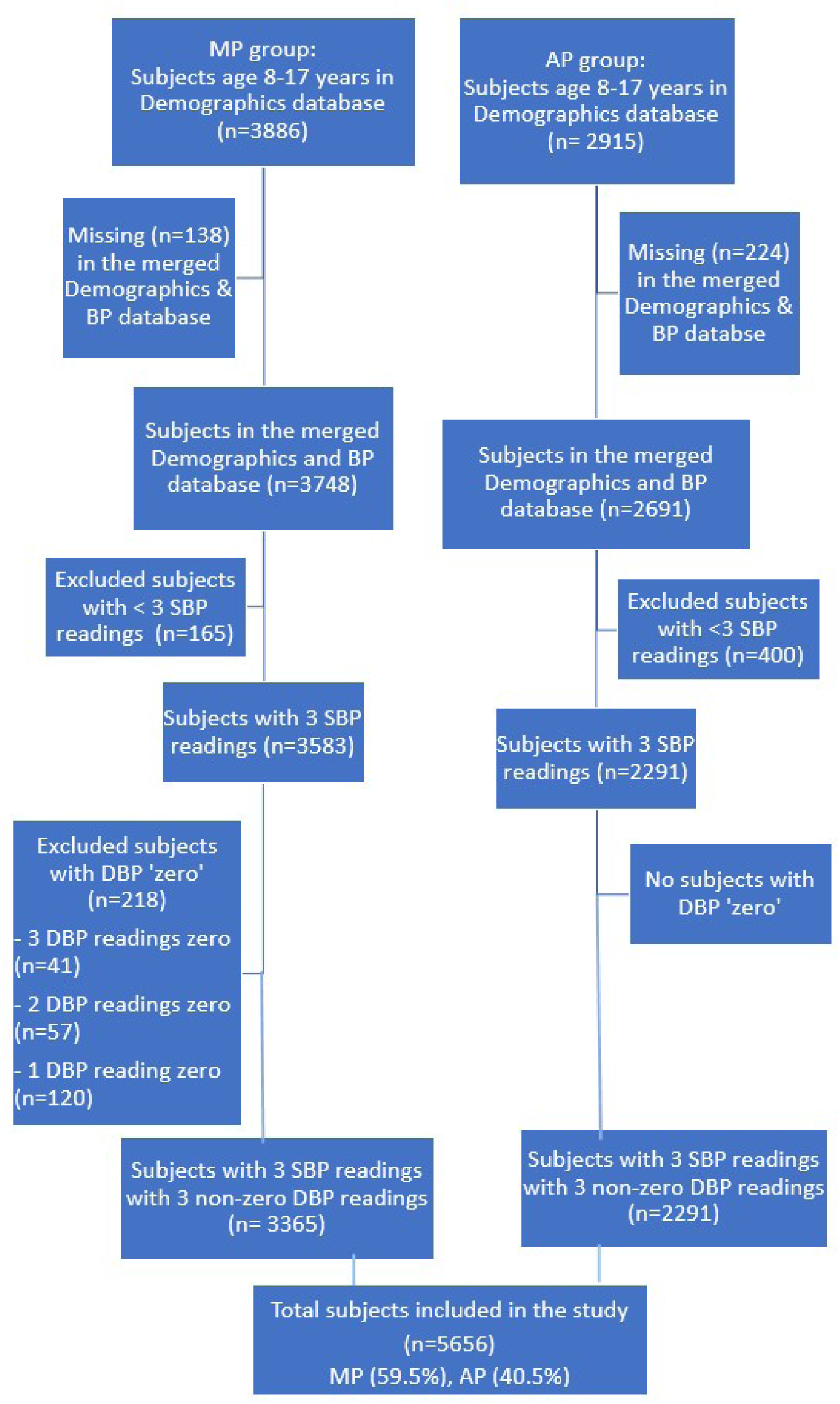
Consort flow diagram demonstrating selection of eligible participants included in the primary analysis among NHANES participants contributing blood pressure data.

Table 1 describes the basic characteristics of subjects. The MP group had a higher percentage of Hispanic white subjects and a lower percentage of participants in the other racial or ethnic groups. Overweight/obesity was noted in 40.6% and was similar between the MP and AP groups. Hypertension-range BP was observed in 4.5% of subjects and was similar in the MP and AP groups.

**Table 1:** Characteristics of study participants.

|  | **Overall**<br>**N=5,656** | **MP group**<br>**N=3,365** | **AP group**<br>**N=2,291** | **P-**<br>**value<sup>‡</sup>** |
| --- | --- | --- | --- | --- |
| Age, years | 12.3 (2.9) | 12.3 (2.9) | 12.3 (2.9) | 0.83 |
| 8-12 | 2,996 (53%) | 1,778 (52.8%) | 1,218 (53.2%) | 0.81 |
| 13-17 | 2,660 (47%) | 1,587 (47.2%) | 1,073 (46.8%) |  |
| Sex |  |  |  | 0.83 |
| Male | 2,829 (50.0%) | 1,687 (50.1%) | 1,142 (49.9%) |  |
| Female | 2,827 (50.0%) | 1,678 (49.9%) | 1,149 (50.2%) |  |
| Race/Ethnicity |  |  |  | **<0.001** |
| Non-Hispanic white | 1,650 (29.2%) | 945 (28.1%) | 705 (30.8%) |  |
| Non-Hispanic Black | 1,377 (24.4%) | 770 (22.9%) | 607 (26.5%) |  |
| Hispanic | 1,697 (30.0%) | 1,130 (33.6%) | 567 (24.8%) |  |
| Other | 932 (16.5%) | 520 (15.5%) | 412 (18.0%) |  |
| BMI Category* |  |  |  | 0.08 |
| Underweight/healthy weight | 3,344 (59.4%) | 2,021 (60.4%) | 1,323 (57.8%) |  |
| Overweight/obesity | 2,282 (40.6%) | 1,325 (39.6%) | 957 (41.8%) |  |
| Hypertension-range BP<sup>†</sup><br>(SBP<sub>1+2+3</sub>/DBP<sub>1+2+3</sub>) | 251 (4.5%) | 143 (4.3%) | 108 (4.7%) | 0.4 |

Mean (SD), or N (column %)
MP: manual protocol (BP was measured using auscultation)
AP: automated protocol (BP was measured using oscillometry)
\*N=30 subjects had missing weight and/or height measurements, hence BMI could not be determined.
<sup>†</sup>N=14 subjects aged <13 years had no height measurements and BP percentiles could not be calculated.
<sup>‡</sup>P-values are calculated by Wilcoxon rank-sum test for continuous data and Fisher's exact and/or Chi-squared tests for categorical data.

Individual-level non-directional BPV analysis showed intra-individual ΔSBP median of 4.0 mmHg and was slightly lower in the MP than AP group (Table 2). The median intra-individual ΔDBP was 6 mmHg and was slightly higher in the MP group. BP SD and VC were low except for the median VC for DBP which was 6.2% for MP group compared to 4.1% for AP group. An intra-individual SBP difference of *at least* 5 mmHg was noted in 49.1% subjects overall (Table 2) and was *less* common in the MP than AP group (45.4% versus 54.8%). Intra-individual ΔDBP ≥5mmHg was seen in 60.7% and was *more* common in the MP than AP group (65.3% versus 54.0%). Notably, intra-individual ΔDBP ≥10 mmHg was twice as common in the MP than AP group (33.3% versus 14.9%). Intra-individual ΔSBP ≥20 mmHg (excessive BPV) was seen in only 0.3% of the MP group as compared to 2.8% of the AP group (Figure 2). In contrast, ΔDBP ≥20 mmHg was higher in the MP than AP group (5.3% versus 3.1%). The maximum ΔSBP was 26 mmHg and 64 mmHg in the MP and AP groups respectively (Tables S3-4), as the subject with ΔSBP 80 mmHg in the AP group seemed to be an outlier (Fig S1). We used ‘distance-based measure’ to define outlier.^38^ This subject (sequence number 120622, Table S4) had sequential SBP readings of 161, 81, and 84 mmHg. The initial SBP seems discordant but may also be from an exaggerated WCE. The highest ΔDBP was 46 mmHg and 53 mmHg for the MP and AP groups respectively (Tables S5-6) because eleven (three in the MP group, eight in the AP group) appeared to be outliers (Fig S2). A review of the subjects with outlier ΔDBP showed the discordant DBP to be the first, second, or third reading in equal proportion in six AP group subjects (Tables S5-6), while the remaining five subjects had no clear single discordant value. In subjects with non-outlier ΔBP ≥ 20 mmHg, the discordant value could be the first, second or third (Tables S3-6).

**Table 2:** BP variability, non-directional (individual-level)

|  | Overall<br>N=5,656 | MP group<br>N=3,365 | AP group<br>N=2,291 | P-Value |
| --- | --- | --- | --- | --- |
| **Ordinal Outcomes** |  |  |  |  |
| ΔSBP (mmHg) | | | | **<0.001** |
| 0-4 | 2,877 (50.9%) | 1,840 (54.7%) | 1,037 (45.3%) |  |
| 5-9 | 2,021 (35.7%) | 1,139 (33.9%) | 882 (38.5%) |  |
| 10-14 | 582 (10.3%) | 342 (10.2%) | 240 (10.5%) |  |
| 15-19 | 103 (1.8%) | 35 (1.0%) | 68 (3.0%) |  |
| $\geq 20$ | 73 (1.3%) | 9 (0.3%) | 64 (2.8%) | |
| ΔDBP (mmHg) | | | | **<0.001** |
| 0-4 | 2,220 (39.3%) | 1,167 (34.7%) | 1,053 (46.0%) |  |
| 5-9 | 1,974 (34.9%) | 1,078 (32.0%) | 896 (39.1%) |  |
| 10-14 | 949 (16.8%) | 738 (21.9%) | 211 (9.2%) |  |
| 15-19 | 265 (4.7%) | 205 (6.1%) | 60 (2.6%) |  |
| $\geq 20$ | 248 (4.4%) | 177 (5.3%) | 71 (3.1%) | |
| **Continuous Outcomes** |  |  |  |  |
| *ΔSBP, mmHg | 5.7 (4.6)<br>4.0 (3.0, 7.0) | 5.2 (3.3)<br>4.0 (2.0, 6.0) | 6.3 (5.9)<br>5.0 (3.0, 8.0) | **<0.001** |
| <sup>†</sup>ΔDBP, mmHg | 7.5 (6.5)<br>6.0 (4.0, 10.0) | 8.1 (6.0)<br>6.0 (4.0, 10.0) | 6.4 (7.0)<br>5.0 (3.0, 8.0) | **<0.001** |
| <sup>‡</sup>SBP SD, mmHg | 3.0 (2.4)<br>2.3 (1.5, 3.8) | 2.8 (1.7)<br>2.3 (1.2, 3.5) | 3.4 (3.2)<br>2.7 (1.5, 4.0) | **<0.001** |
| <sup>§</sup>DBP SD, mmHg | 3.9 (3.4)<br>3.1 (2.0, 5.0) | 4.3 (3.1)<br>3.5 (2.0, 5.8) | 3.4 (3.8)<br>2.5 (1.5, 4.0) | **<0.001** |
| SBP VC (%) | 2.9 (2.4)<br>2.4 (1.4, 3.6) | 2.6 (1.7)<br>2.3 (1.2, 3.5) | 3.2 (3.2)<br>2.5 (1.5, 3.9) | **<0.001** |
| <sup>#</sup>DBP VC (%) | 7.0 (6.8)<br>5.2 (3.1, 8.6) | 8.1 (7.4)<br>6.2 (3.7, 10.1) | 5.5 (5.5)<br>4.1 (2.5, 6.4) | **<0.001** |

Mean (SD), Median (IQR) or N (column %)
MP: manual protocol (BP was measured using auscultation)
AP: automated protocol (BP was measured using oscillometry)
\* ΔSBP, <sup>†</sup> ΔDBP = the difference between the maximum and minimum SBP and DBP value for each subject respectively.
<sup>‡</sup>SBP SD, <sup>§</sup>DBP SD: standard deviation of the 3 SBP readings and 3 DBP readings for each subject respectively.
<sup>||</sup>SBP VC, <sup>#</sup>DBP VC: BP variability coefficient of the 3 BP readings for each subject respectively. (SD/mean SBP) x 100. (SD/mean DBP) x 100.
\*P-values are calculated by Wilcoxon rank-sum test for continuous data and Fisher's exact and/or Chi-squared tests for categorical data.

**Figure 2:**
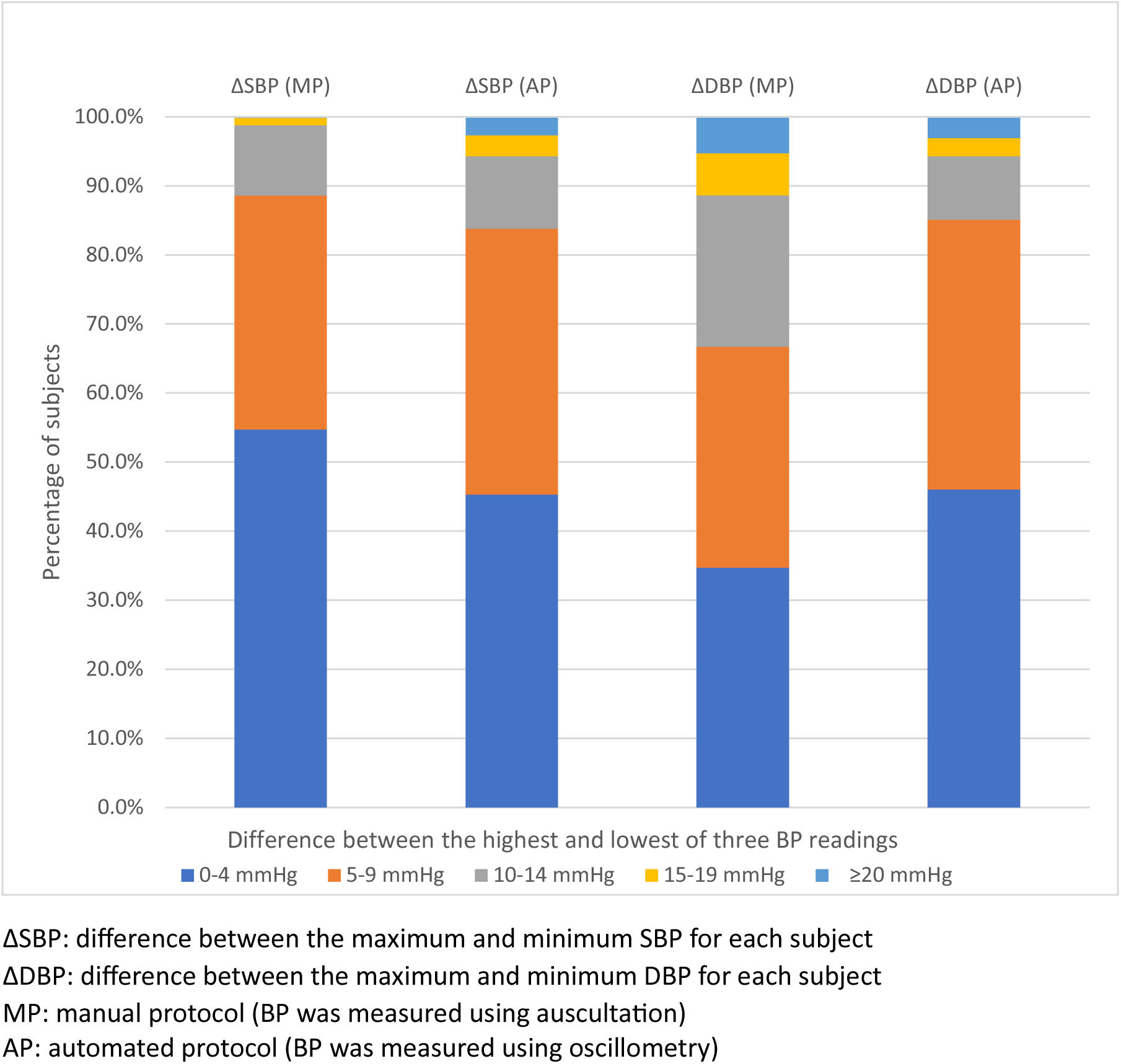
Distribution of ΔSBP and ΔDBP (individual-level)

Individual-level directional BPV measures included ARV (Table 3). The median SBP ARV was 3.0 mmHg, being similar in the MP and AP groups, while median DBP ARV was 4.0 mmHg and slightly higher in the MP than AP group. Notably, a DBP ARV ≥10 mmHg was twice as frequent in the MP group versus AP group (12.9% versus 5.8%). Individual-level directional BPV was also analyzed by the highest of three BP readings. While the first reading was the highest of all three BP readings most often (44.2% and 42.4% for SBP and DBP respectively), in over half the subjects the second or third readings were the highest in both the MP and AP groups (Table 3). At the individual level, a consistent pattern for decreasing BP (of at least 1 mmHg) was seen in 12.2% and 14.2% of subjects for SBP and DBP respectively and was more common in the AP than MP group for both SBP and DBP (Table 3). The opposite trend of consistent increase in individual-level BP readings was noted in 10.3% and 12.0% of subjects for SBP and DBP respectively and was more often seen in the AP group for SBP.

**Table 3:** BP variability, directional (individual level)

|  | Overall<br>N=5,656 | MP group<br>N=3,365 | AP group<br>N=2,291 | P-value<sup>†</sup> |
| --- | --- | --- | --- | --- |
| **Continuous Outcomes** |  |  |  |  |
| ARV* SBP, mmHg | 3.7 (3.2)<br>3.0 (2.0, 5.0) | 3.4 (2.4)<br>3.0 (2.0, 5.0) | 4.1 (4.0)<br>3.0 (2.0, 5.0) | **<0.001** |
| ARV DBP, mmHg | 4.9 (4.5)<br>4.0 (2.0, 6.0) | 5.3 (4.2)<br>4.0 (3.0, 7.0) | 4.2 (4.9)<br>3.0 (2.0, 5.0) | **<0.001** |
| **Discrete Outcomes** |  |  |  |  |
| ARV SBP (mmHg) |  |  |  | **<0.001** |
| 0-4 | 4,153 (73.4%) | 2,515 (74.7%) | 1,638 (71.5%) |  |
| 5-9 | 1,290 (22.8%) | 775 (23.0%) | 515 (22.5%) |  |
| 10-14 | 158 (2.8%) | 71 (2.1%) | 87 (3.8%) |  |
| 15-19 | 29 (0.5%) | 3 (0.1%) | 26 (1.1%) |  |
| ≥ 20 | 26 (0.5%) | 1 (0.03%) | 25 (1.1%) |  |
| ARV DBP (mmHg) |  |  |  | **<0.001** |
| 0-4 | 3,498 (61.8%) | 1,807 (53.7%) | 1,691 (73.8%) |  |
| 5-9 | 1,592 (28.2%) | 1,125 (33.4%) | 467 (20.4%) |  |
| 10-14 | 387 (6.8%) | 317 (9.4%) | 70 (3.1%) |  |
| 15-19 | 100 (1.8%) | 74 (2.2%) | 26 (1.1%) |  |
| ≥ 20 | 79 (1.4%) | 42 (1.3%) | 37 (1.6%) |  |
| Highest SBP |  |  |  | **0.001** |
| SBP<sub>1</sub> | 2,499 (44.2%) | 1,548 (46.0%) | 951 (41.5%) |  |
| SBP<sub>2</sub> | 1,722 (30.5%) | 1,012 (30.1%) | 710 (31.0%) |  |
| SBP<sub>3</sub> | 1,435 (25.4%) | 805 (23.9%) | 630 (27.5%) |  |
| Highest DBP |  |  |  | **0.047** |
| DBP<sub>1</sub> | 2,400 (42.4%) | 1,383 (41.1%) | 1,017 (44.4%) |  |
| DBP<sub>2</sub> | 1,687 (29.8%) | 1,023 (30.4%) | 664 (29.0%) |  |
| DBP<sub>3</sub> | 1,569 (27.7%) | 959 (28.5%) | 610 (26.6%) |  |
| Decreasing throughout |  |  |  |  |
| SBP | 692 (12.2%) | 357 (10.6%) | 335 (14.6%) | **<0.001** |
| DBP | 803 (14.2%) | 434 (12.9%) | 369 (16.1%) | **0.001** |
| Increasing throughout |  |  |  |  |
| SBP | 582 (10.3%) | 300 (8.9%) | 282 (12.3%) | **<0.001** |
| DBP | 676 (12.0%) | 425 (12.6%) | 251 (11.0%) | 0.06 |

Mean (SD), Median (IQR) or N (column %)
MP: manual protocol (BP was measured using auscultation)
AP: automated protocol (BP was measured using oscillometry)
\*ARV = Average real variability (see text).
SBP<sub>1</sub> = first SBP reading; SBP<sub>2</sub> = second SBP reading; SBP<sub>3</sub> = third SBP reading; DBP<sub>1</sub> = first DBP reading; DBP<sub>2</sub> = second DBP reading; DBP<sub>3</sub> = third DBP reading
<sup>†</sup>P-values are calculated by Wilcoxon sum-rank tests for continuous data and Fisher's exact and/or Chi-squared tests for categorical data.

Individual-level directional BPV was also assessed by comparing each subject’s first BP reading with subsequent BP and total BP respectively (Table S2). The median *absolute* difference between initial and subsequent SBP was 2.5 mmHg and was slightly lower in the MP than AP group. The median *absolute* difference between each subject’s initial and subsequent DBP was 3.0 mmHg and was higher in the MP than AP group. Similar relationship was noted between initial and total SBP and DBP, but the differences were smaller. Notably, a ΔSBP_sub_ ≥ 5 mmHg was seen in 24.9% of subjects, being *higher* in 13.4% and *lower* in 11.5% (Figure 3) with no difference between the MP and AP groups (Table S2). A ΔDBP_sub_ ≥5 mmHg was seen in 34.5% of subjects, with 18.2% of subjects having *higher* DBP_1_ than subsequent DBP (Figure 3), which was noted more often in the MP than AP group (Table S2). Similar but smaller and less frequent differences were seen between individual-level initial and total BP.

**Figure 3:**
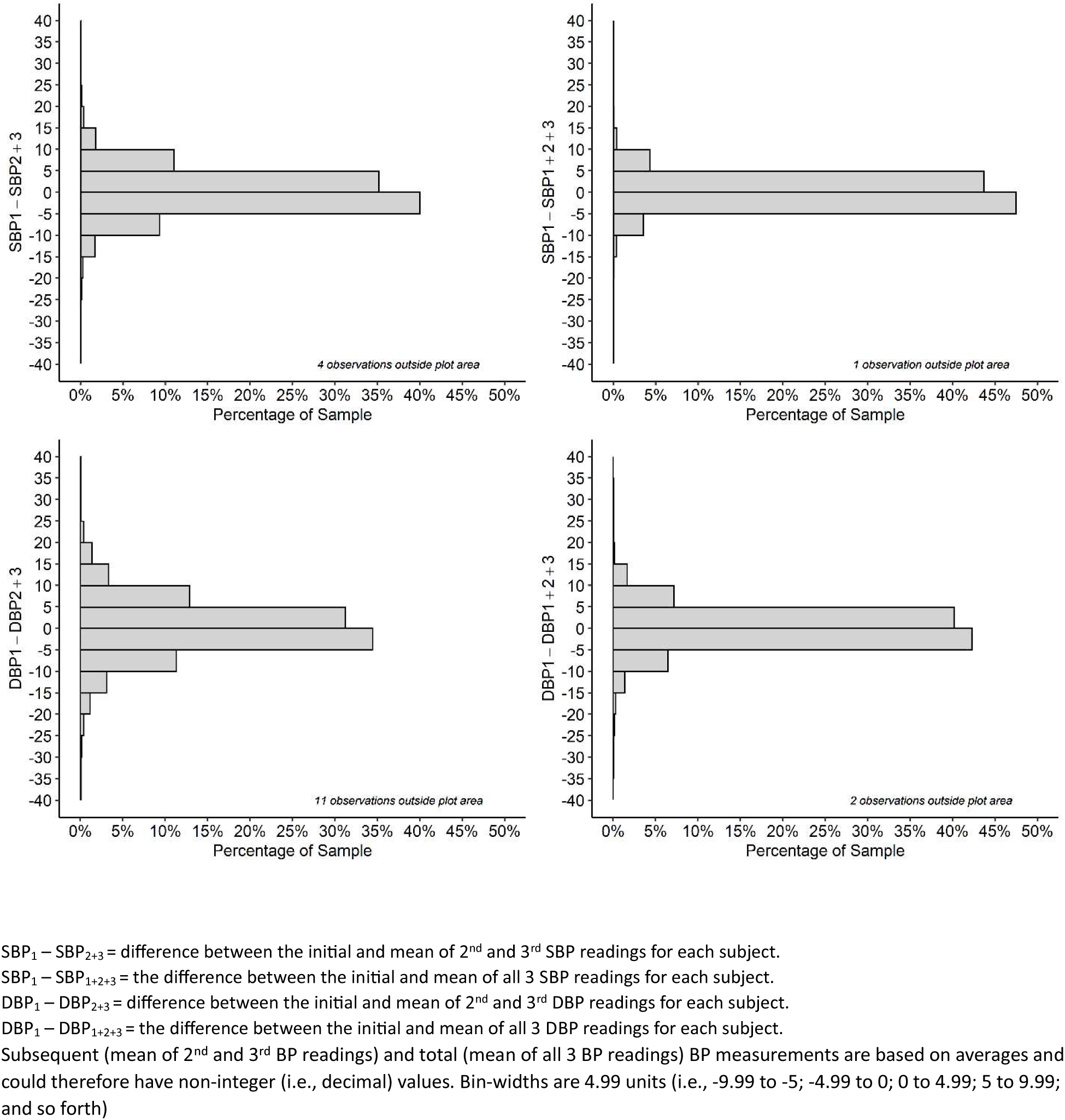
Difference between initial and subsequent and total BP (individual-level)

Group-level directional BPV analysis (Table S1) showed that the mean SBP decreased from the first to third reading in both the MP and AP groups as did mean DBP in the AP group. However, in the MP group, mean DBP decreased from the first to second but went up with the third reading. Additionally, group-level mean SBP_1_ and DBP_1_ were higher than subsequent and total BP except in the MP group where DBP_1_ was similar to total DBP. Group-wide subsequent BP was mostly lower than total BP or similar.

As higher DBPV (diastolic blood pressure variability) was a finding of our study, we assessed subjects with DBP ‘zero’ although they were excluded from the primary analysis. A DBP of ‘zero’ in one or more of the BP readings was noted exclusively in the MP group and occurred in 6.1% (218/3583). Most of these subjects (55.1%) had a single DBP ‘zero’, followed by subjects with two ‘zero’ DBP readings (26.1%) and all three ‘zero’ DBP readings in 18.8%.

## DISCUSSION

Our study in a diverse community-based group of children and adolescents participating in NHANES shows significant within-visit BPV among three readings. About half the subjects had a difference of ≥5 mmHg among the three BP readings and some had excessive discrepancy with a difference of 20 mmHg or more. A quarter of subjects had a difference of ≥5 mmHg between the initial and subsequent BP. Contrary to a general assumption that BP decreases with repeat measurements, we found that BP may increase across subsequent measurements, and there is no consistent pattern in most subjects. In comparing the MP and AP groups, SBPV (systolic blood pressure variability) was slightly higher in the AP group while DBPV was greater in the MP group.

We noted ΔBP of < 5mmHg for SBP and DBP in 50.9% and 39.3% of all subjects, respectively. A study in Australian children with two automated BP readings also found 50% of subjects with ΔSBP -4 to 4 mmHg; ΔDBP was not reported.^23^ Studies in adults show similar variability in SBP but lower variability in DBP than our study. In adults with three manual BP readings, 57.4% and 68.3% of subjects had ΔSBP and ΔDBP 0–5 mmHg respectively as compared to 54.7% and 34.7% in our MP group.^39^ Two adult studies of within-visit BPV with three manual BP readings found ΔBP ≥6 mmHg for SBP in 42.6-64.5% but only in 31.7-32.3% subjects for DBP.^39,40^ In a similar trend of higher DBPV in children, we noted ΔBP ≥10 mmHg for SBP and DBP in 13.4% and 26% of our subjects respectively compared to 12.9% and 13% in adult NHANES data (n=53,737).^41^ A 1987 study of manual BP readings in children echoes our findings of higher DBPV than SBPV and also noted that BPV in children is considerably higher than BPV in adults.^42^

The finding of high DBPV in our study despite excluding subjects with DBP ‘zero’ maybe related to subjects with manual DBP as low as 2-14 mmHg although automated DBP readings were not as low (Tables S5,6). Our rate of DBP of ‘zero’ in the MP group was similar to the Bogalusa Heart Study (6.1% vs. 7.4%).^32^ The reason for higher DBPV in children than adults are not clear but may be related to ‘wellness’ factors such as age, lifestyle differences and lower hypertension prevalence. A pediatric study noted that younger age and lower BP were associated with higher DBPV.^42^ In an adult study, within-visit DBPV was positively associated with physical activity and inversely related with hypertension diagnosis.^43^ Interestingly, SBPV ≥ 10 mmHg was associated with diabetes status progression, but DBPV ≥ 10 mmHg was not. In children with untreated primary hypertension, SBPV was higher than DBPV and family history of hypertension was associated with lower DBPV.^44^ Higher DBPV than SBPV in our study may be related to most of our subjects being normotensive. Higher sympathetic transduction was associated with greater DBPV in young healthy adults, however available pediatric data suggest that sympathetic nervous system activity decreases from birth to adolescence.^45,46^ The role of arterial stiffness is unclear; a study in China noted a temporal relationship between long-term BPV and arterial stiffness^47^, however a study in France found no association between short-term BPV and arterial stiffness.^48^

In our study, excessive BPV (ΔBP ≥ 20 mmHg) rate was much lower for SBP than DBP in the MP group (0.3% vs. 5.3%) while AP group had similar rate (2.8% vs. 3.1%), likely from the protocolized calculation. Adult data for three *manual* BP readings show a reverse trend with higher frequency of ΔSBP ≥20 mmHg (1.3%-1.7%) but only 0.5% for ΔDBP ≥20 mmHg.^39,40^ The phenomenon of lower SBPV and higher DBPV in children than in adults was noted in a study with manual BP readings.^42^ We found maximum ΔBP of 26 mmHg, 64 mmHg, 46 mmHg, and 53 mmHg for manual SBP, automated SBP, manual DBP, and automated DBP respectively. Although these appear excessively high, other studies have described similar discrepancy in intra-individual BP readings with reports of ΔSBP 50 mmHg and ΔDBP 40 mmHg with a full urinary bladder^49^, ΔSBP 52 mmHg^40^, and ΔSBP 75 mmHg with ΔDBP 36 mmHg related to alerting reaction^50^. Currently, it is unclear what degree of BPV is excessive and should be discarded.

Accordingly, arbitrary thresholds appear to be in use. In the Australian Health Survey, a difference >10 mmHg between the first two BP readings triggered a repeat BP measurement, while in the Canadian Health Measures Survey, based on pre-test data, ΔSBP ≥30 mmHg or ΔDBP ≥20 mmHg prompted a repeat BP measurement.^23,51^ The NHANES data protocol is strict but does not specify BPV limits. Ambulatory blood pressure monitoring (ABPM) protocols set high and low parameters (SBP >260 mmHg or <70 mmHg, and DBP >150 mmHg or <40 mmHg are discarded) but do not discard readings with excessive differences.^37^

In our study, first BP was not always the highest, in contrast to other studies that describe a consistent decrease with repeat BP measurements however these studies were smaller and mostly compared group rather than individual BP readings.^8,51–53^ We found that the highest of the three readings was the first, second, and third reading in 45%, 30%, and 25% of subjects; corresponding numbers in an adult study were 54%, 25%, and 21% respectively.^54^ We noted ΔSBP_sub_ ≥5 mmHg in 24.9% of all subjects and among these, subsequent BP was higher in 13.4% and lower in 11.5% subjects. The trend for subsequent BP to be lower or higher was also noted in a study in Australian children where the proportion of subjects with a decrease, increase, or no change from first to second automated SBP was 58%, 32%, and 10%.^23^ We noted that individual-level BP from the first to third reading showed a consistent decrease, a consistent increase, and a fluctuating pattern in about 13%, 12%, and 75% of subjects respectively with a comparable trend in an adult study with three automated BP readings with corresponding rates of 29%, 7%, and 64% respectively.^54^ This trend in our study may be explained by regression to the mean (or regression dilution bias) described in another NHANES study whereby low initial BP values tended to rise and higher values to fall.^55^

The significance of BPV includes impact on health, hypertension diagnosis, and subsequent treatment. In adults, there are mixed data about the association of high within-visit BPV with cardiovascular health risks such as stroke and diabetes^43,56,57^, and the role of short-term BPV in hypertension misdiagnosis is being recognized.^58,59^ In children, there are emerging, mostly cross-sectional, data on the association of BPVW with health. While some studies in children shows associations between short-term BPV derived from ABPM with cardiac remodeling features such as LVMI^60^ and relative wall thickness^61^, another study found no correlation.^62^ In hospitalized children, BPV was associated with acute kidney injury.^63^ In children with chronic kidney disease, ABPM-derived BPV was associated with hypertension but not cognitive abnormalities.^64,65^

### Perspectives

Our study in a large group of healthy children found that there is considerable variability within three BP readings, both with manual and automated BP methodology. Our results also challenge the widely held belief that BP decreases with repeat measurements. Only a small minority of subjects had a consistent decrease in BP with repeat measurements and a consistent *increase* in BP was seen in some subjects. In the vast majority, there was no consistent trend of increase or decrease among the three BP readings. Our findings suggest that single BP readings may not represent a child’s BP status but future studies that include target organ damage will clarify this further.

### Study limitations

Our study of BPV had some limitations. First, our study represents a series of cross-sectional analyses. Second, the NHANES BP protocol is rigorous while methodological errors are common in routine clinical care and may affect applicability of our findings to the general population. Third, we grouped participants with BP in the hypertension range and normotensive range together for analysis and did not have information about hypertension diagnosis or use of anti-hypertension medications. Fourth, we compared BPV in manual BP readings with automated BP readings, but these were not obtained in the same subjects. Although we did not exclude subjects with hypertension, the proportion with BP in the hypertension range was relatively low (∼4%). Although real-life clinical practice may not allow the rigor adopted by NHANES, our study findings may persuade attention to appropriate BP measurement protocol as much as possible.

## CONCLUSIONS

In conclusion, there is significant within-visit BPV with three BP readings in healthy children. DBPV tends to be higher in the MP than AP groups while SBPV is slightly higher in AP than MP group. BP can vary by as much as 20 mmHg among three readings and the discordant reading may be the first, second, or third, which challenges the paradigm that the first reading is always the highest and should be discarded. BP may consistently decrease or increase with three BP readings however in the majority, there is no consistent trend. Over half the subjects have the second or third reading as their highest. Our findings may impact the diagnosis of hypertension especially in younger children in whom the difference between normal and hypertension threshold is about 4 mmHg for SBP and about 3 mmHg for DBP. Longitudinal studies are required to clarify the health implications of BPV and number of required readings in youth. Information about the number of BP readings obtained should be available in pediatric hypertension trials.

### Novelty and Relevance

#### What Is New?

- BP can vary by as much as 20 mmHg or more between the highest and lowest of three BP readings especially for diastolic BP.
- A difference of at least 5 mmHg between initial BP and the average of second and third readings was noted in 25% and 35% of all subjects for SBP and DBP respectively, being higher and lower in similar proportion.
- BP variability is seen with both manual and automated BP readings and is mostly similar however diastolic BP is more variable with manual than automated BP readings.
- Pediatric SBPV (systolic blood pressure variability) seems to be similar to adults, but DBPV is higher.

#### What Is Relevant?

- BP may increase or decrease with three measurements at a single encounter.

#### Clinical/Pathophysiological Implications?

- A single BP reading may not be representative of BP status which has implications in the diagnosis and management of pediatric hypertension.
- Current recommendations to obtain single BP reading may need to be re-evaluated.

## Data Availability

Included

## Non-standard Abbreviations and Acronyms

ABPM: ambulatory blood pressure monitoring
AP: automated protocol
ARV: average real variability
BMI: body mass index
BP: blood pressure
BPV: blood pressure variability
DBP: diastolic blood pressure
DBPV: diastolic blood pressure variability
MP: manual protocol
NHANES: National Health and Nutrition Examination Survey
SD: standard deviation
SBP: systolic blood pressure
SBPV: systolic blood pressure variability
VC: variability coefficient

## Acknowledgments, Sources of Funding, & Disclosures

### Acknowledgments

We are grateful for help with data collection provided by Dr. Brian Lee, PhD, MPH (Statistician at Children’s Mercy Hospital, 2401 Gillham Rd Kansas City, MO 64108). Dr. Lee has seen and approved mention of his name in the article.

## Supplemental Material

Figures S1, S2

Tables S1 to S6

